# Investigation of HLA susceptibility alleles and genotypes with hematological disease among Chinese Han population

**DOI:** 10.1101/2023.01.31.23285238

**Authors:** Ye-Mo Li, Yu-Xia Li, Dai-Yang Li, Lin An, Zhi-Yang Yuan, Zhong-Liang Liu, Ke-Ming Du, Zhong-Zheng Zheng

## Abstract

**Background:** Serveral genes involved in the pathogenesis have been identified and a crucial role is known to be played by the human leukocyte antigen (HLA) system. However, the relationship between HLA and a cluster of hematological diseases has been rarely reported in China.

**Methods:** Blood samples (n=123,913) from 43,568 patients and 80,345 individuals without known pathology were genotyped using sequence-based typing (SBT) for HLA class I and II.

**Results:** The HLA-A*11:01, B*40:01, C*01:02, DQB1*03:01, and DRB1*09:01 were common in China. Additionally, compared to controls, in malignant hematologic diseases, 3 high-frequency alleles (DQB1*03:01, DQB1*06:02, and DRB1*15:01) were risky. And we observed 7 high-frequency risk alleles (A*01:01, B*46:01, C*01:02, DQB1*03:03, DQB1*05:02, DRB1*09:01, and DRB1*14:54) and 8 high-frequency susceptible genotypes (A*11:01-A*11:01, B*46:01-B*58:01, B*46:01-B*46:01, C*01:02-C*03:04, DQB1*03:01-DQB1*05:02, DQB1*03:03-DQB1*06:01, DRB1*09:01-DRB1*15:01, and DRB1*14:54-DRB1*15:01) for benign hematologic diseases.

**Conclusion:** Our results support the association between HLA alleles/genotypes and multiple hematological disorders, which is essential in disease surveillance.

## 1 INTRODUCTION

Hematological diseases are becoming a global public health problem. Hematological disorders comprise a large number of malignant and benign hematologic manifestations, including leukocyte abnormalities, erythrocyte disorders, hemorrhagic disease, and myeloproliferative disorders. It is well-known that blood diseases often lead to shortened lifespans and poor quality of life. With the development of gene therapy, previous studies have confirmed that genetic factors influence the treatment of blood disorders**(Goyal *et al*. 2022)**. As a result, the research of disease-susceptibility genes might promote the clinical formulation of reasonable treatment plans.

Human leukocyte antigen (HLA) is grouped into three subclasses regions: class I (HLA-A, HLA-B, and HLA-C, *etc*.), class II (HLA-DQ, HLA-DR, HLA-DP, *etc*.), and class III containing genes implicated in inflammatory responses, leukocyte maturation, and the complement cascade**(Dendrou *et al*. 2018)**. HLA is highly polymorphic, and alleles/genotypes have been reported to be associated with over 500 diseases**(Zhong *et al*. 2019; Ebrahimi *et al*. 2020; Roerden *et al*. 2020; Zaimoku *et al*. 2021)**. HLA genes play a pivotal role in successful hematopoietic stem cell transplantation (HSCT)**(Gragert *et al*. 2014)**. What’s more, HLA polymorphism has been shown to be associated with prognosis in hematological patients**(Shouval *et al*. 2019)**. Undoubtedly, due to the distribution of alleles/genotypes in the HLA system varying between different populations, it is critical to observe independent evidence on the association between HLA and hematological diseases in China.

In the present study, we targeted to investigate alleles and genotypes at five HLA loci (-A, -B, -C, -DRB1, and -DQB1) hematologic patients compared with individuals without known pathology in China. Importantly, we paid close attention to the association of HLA class I and II with malignant and benign hematologic diseases. This will be followed by the distribution of HLA alleles/genotypes and the presentation of how disease-associated genes are defined.

## 2 MATERIALS AND METHODS

### 2.1 Study population

From June 2012 to July 2021, blood samples were collected from Han population from over 600 hospitals in 29 provinces across China. All subjects received written informed consent for the study. This study was approved by the Ethics Committee of Shanghai Tissuebank Medical Laboratory.

Herein, the malignant hematologic diseases included acute lymphatic leukemia (ALL), chronic myeloid leukemia (CML), acute myeloid leukemia (AML), myelodysplastic syndromes (MDS), and lymphoma. The benign hematologic diseases in this study were composed of hemophagocytic lymphohistiocytosis (HLH), aplastic anemia (AA), and thalassemia. Leukemia diagnosis followed the French-American-British (FAB) classification. Diagnosis of other disease types was according to previously reported criteria.

A total of 123,913 samples, including 43,568 patients and 80,345 individuals without known pathology. There were 19,354 patients with precise diagnostic information, of whom 10,443 had been diagnosed with malignant hematologic diseases (3,422 of ALL, 4,681 of AML, 923 of CML, and 1,417 of MDS). Patients with benign hematologic diseases consisted of 415 with HLH, 2,868 with AA, and 535 with thalassemia. There were 790 cases of lymphoma and 4,303 cases of other diseases (**Fig S1**).

### 2.2 DNA extraction

Blood samples were collected from patients and controls. Genomic DNA was extracted from peripheral blood using the QIARamp Blood Kit (Qiagen, Hilden, Germany). HLA class I (-A, -B, and -C) and II (mainly, -DRB1 and -DQB1) alleles were subtyped using primer pairs recommended by the International Histocompatibility Working Group (IHWG)(Petersdorf *et al*. 2013). High-resolution genotyping of HLA groups was performed using the sequence-based typing (SBT) method by the HLA Genotyping Kit (Shanghai Tissuebank Biotech, Shanghai, China).

The reaction conditions were as follows: pre-denaturation at 96°C for 2 min, denaturation at 96°C for 15 s, annealing at 65°C for 15 s, extension at 72°C for 1 min, 40 cycles, extension at 72°C for 7 min, and storage at 4°C. The reaction product was stored at 4°C. 5 μl of the amplification product was extracted by 1.5% agarose gel electrophoresis and the results were observed under UV light.

### 2.3 Statistical analysis

Allele frequency was calculated by referring to the Chinese common and well-documented (CWD) Allele Table(He *et al*. 2018) as follows: allele frequency = number of genes/(total number of a disease*2), genotype frequency = number of genotypes/total number of diseases. HLA allele frequency and genotype were determined by the direct counting method.

All analyses were performed using R version 4.1.0. A Chi-square test was applied to compare the variability of HLA alleles and genotypes between patients and individuals without known pathology. The prevalence of patients was counted according to diagnostic information, and the association strength was estimated by odds ratio (OR) and 95% confidence interval (CI). Then, Bonferroni’s correction was used to adjust the P-value, which was considered statistically significant when it was less than 0.05.

## 3 RESULTS

### 3.1 Characteristics of HLA alleles frequency

In total, 202 alleles of HLA-A, 282 of HLA-B, 173 of HLA-C, 86 of HLA-DQB1 and 149 of DRB1 were analyzed. As shown in **Fig 1**, alleles A*11:01 (22.91%), A*24:02 (15.61%), A*02:01 (11.63%), A*02:07 (9.30%), and A*33:03 (8.08%) had the highest frequencies at HLA-A locus. B*40:01 (11.62%), B*46:01 (11.22%), B*58:01 (6.31%), B*13:01 (5.74%), and B*51:01 (5.43%) were the top five high-frequency HLA-B alleles. Details and the high-frequency alleles of HLA-C, -DQB1 and -DRB1 were shown in **Fig 1**. Among them, A*11:01, A*02:01, A*02:07, A*33:03, B*40:01, B*46:01, B*58:01, C*01:02, C*07:02, C*06:02, DQB1*05:02, DQB1*06:02, DRB1*15:01, DRB1*12:02 and DRB1*07:01 had significantly differences between patients and controls (Pc < 0.05).

**Figure 1.**
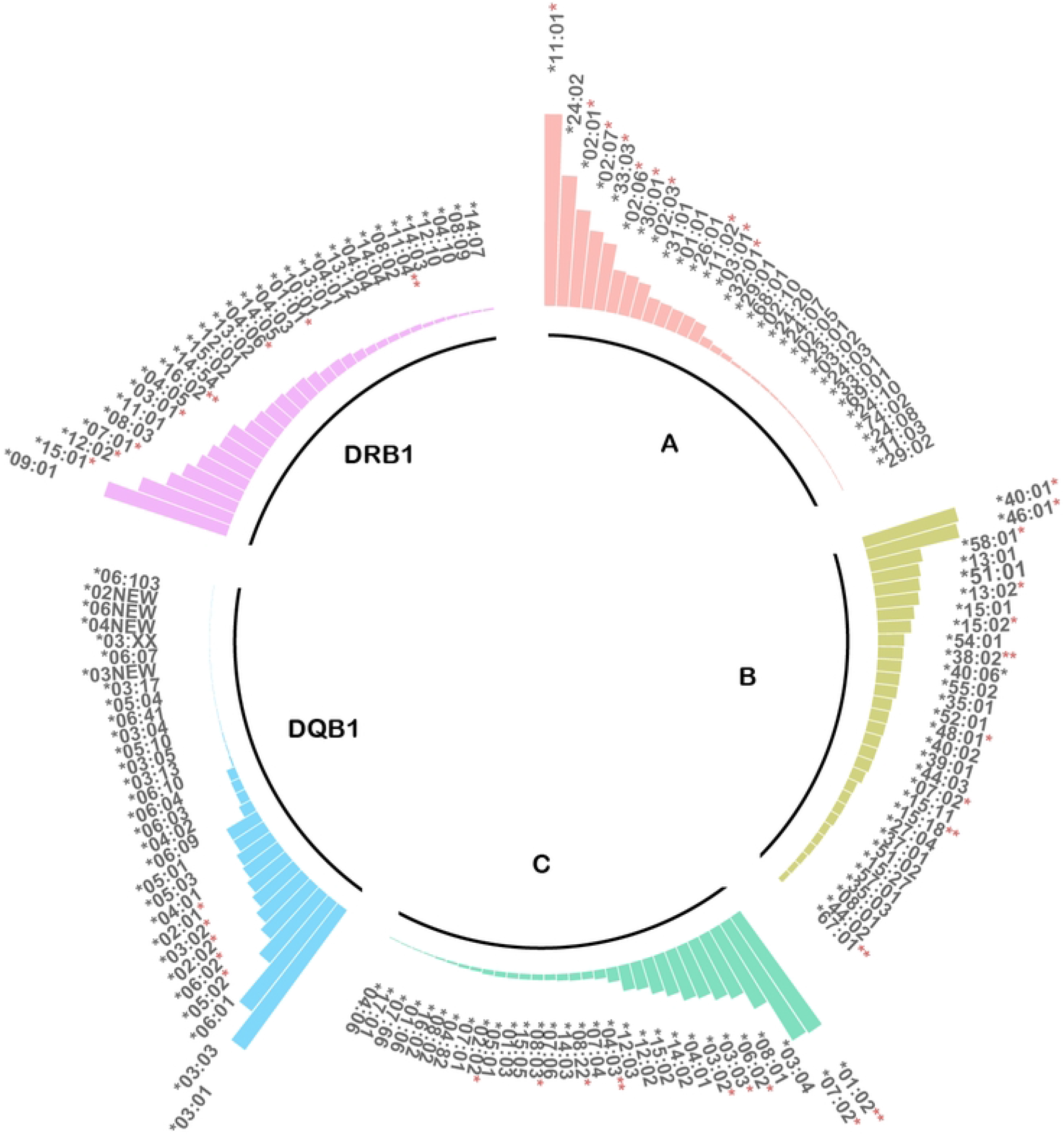
Characteristics of HLA alleles frequency. Frequencies of HLA-A, -B, -C, -DQB1, -DRB1 alleles between patients and control subjects were compared. * Pc < 0.05, ** Pc < 0.01.

### 3.2 Potential protective and risk HLA-A, -B, -C, -DQB1, and -DRB1 alleles and genotypes

#### 3.2.1 Malignant hematologic diseases

As seen in **Table 1**, there were 4 protective and 2 risk alleles in AML: the former included A*11:01 (OR: 0.91; 95% CI: 0.86-0.95, Pc = 0.02), B*13:11 (OR: 0.85; 95% CI: 0.77-0.93, Pc = 0.04), C*03:04 (OR: 0.85; 95% CI: 0.79-0.92, Pc = 0.02), and DQB1*05:02 (OR: 0.82; 95% CI: 0.76-0.89, Pc = 0.02), while the latter included A*03:01 (OR: 1.30; 95% CI: 1.15-1.48, Pc = 0.02) and DQB1*03:01 (OR: 1.09; 95% CI: 1.04-1.15, Pc = 0.05).

**Table 1.** Differences in the most common HLA-A, -B, -C, -DQ, and -DR alleles between patients with malignant hematologic disorders and controls.

| Disease | Allele | Frequency in patients (%) | Frequency in controls (%) | OR (95%CI) | P-value | Pc |
| --- | --- | --- | --- | --- | --- | --- |
| AML | A*11:01 | 21.67 | 23.39 | 0.91<br>(0.86-0.95) | <0.01 | 0.02 |
|  | B*13:01 | 4.92 | 5.76 | 0.91<br>(0.86-0.95) | <0.01 | 0.04 |
|  | C*03:04 | 9.45 | 10.91 | 0.85<br>(0.77-0.93) | <0.01 | 0.02 |
|  | DQB1*03:01 | 22.17 | 20.74 | 1.09<br>(1.04-1.15) | <0.01 | 0.05 |
| MDS | A*11:01 | 19.37 | 23.39 | 0.79<br>(0.72-0.86) | <0.01 | 0.01 |
|  | B*46:01 | 8.05 | 11.45 | 0.68<br>(0.59-0.78) | <0.01 | 0.01 |
|  | C*01:02 | 14.68 | 16.84 | 0.85<br>(0.77-0.94) | <0.01 | 0.02 |
|  | DQB1*06:02 | 10.23 | 7.08 | 1.50<br>(1.32-1.69) | <0.01 | 0.01 |
|  | DRB1*15:01 | 13.62 | 11.53 | 1.21<br>(1.09-1.35) | <0.01 | 0.01 |

In contrast, MDS seems to have more protective and risk factors for this study, corresponding to 11 (including 3 of HLA-A, 2 of HLA-B, 3 of HLA-C, 2 of HLA-DRB1, and 1 of HLA-DQB1) and 14 (including 5 of HLA-B, 3 of HLA-C, 3 of HLA-A, 2 of HLA-DRB1, and 1 of HLA-DQB1) alleles. For high frequency alleles, there were A*11:01 (OR: 0.79; 95% CI: 0.72-0.86, Pc = 0.01), B*46:01 (OR: 0.68; 95% CI: 0.59-0.78, Pc = 0.01), C*01:02 (OR: 0.85; 95% CI: 0.77-0.94, Pc = 0.02). and DQB1*06:02 (OR: 1.50; 95% CI: 1.32-1.69, Pc = 0.01).

#### 3.2.2 Benign hematologic diseases

As shown in **Table 2**, it is clear that anemia had the largest number of alleles with potential protective and risk effects in the present study. Among them, HLH possessed 3 protective alleles [i.e. B*15:02 (OR: 0.43; 95% CI: 0.26-0.72), Pc = 0.02, C*08:01 (OR: 0.61; 95% CI: 0.45-0.83, Pc = 0.02), and DQB1*03:01 (OR: 0.74; 95% CI: 0.62-0.89, Pc = 0.04)] and 4 risk alleles [i.e. A*01:01 (OR: 2.04; 95% CI: 1.50-2.76, Pc = 0.02) and DRB1*10:01 (OR: 2.54; 95% CI: 1.76-3.66, Pc = 0.02)]. Twenty-three alleles expressed protective effects against thalassemia, most of which were at loci A and B (6 and 8, respectively), and the remaining at loci C, DRB1 and DQB1 (3 of each), among which A*02:01 (OR: 0.40; 95% CI: 0.30-0.53, Pc < 0.01) and C*03:03 (OR: 0.62; 95% CI: 0.46-0.84, Pc = 0.01) had the highest frequencies. While B*46:01 (OR: 1.67; 95% CI: 1.43-1.96, Pc < 0.01), DQB1*05:02 (OR: 3.05; 95% CI: 2.64-3.52, Pc < 0.01), and DRB1*14:54 (OR: 2.76; 95% CI: 2.24-3.40, Pc < 0.01) were the highest frequency among 20 risk alleles (A/B/C/DRB1/DQB1 loci correspond to 3, 5, 5, 4 and 3 alleles, respectively).

**Table 2.**
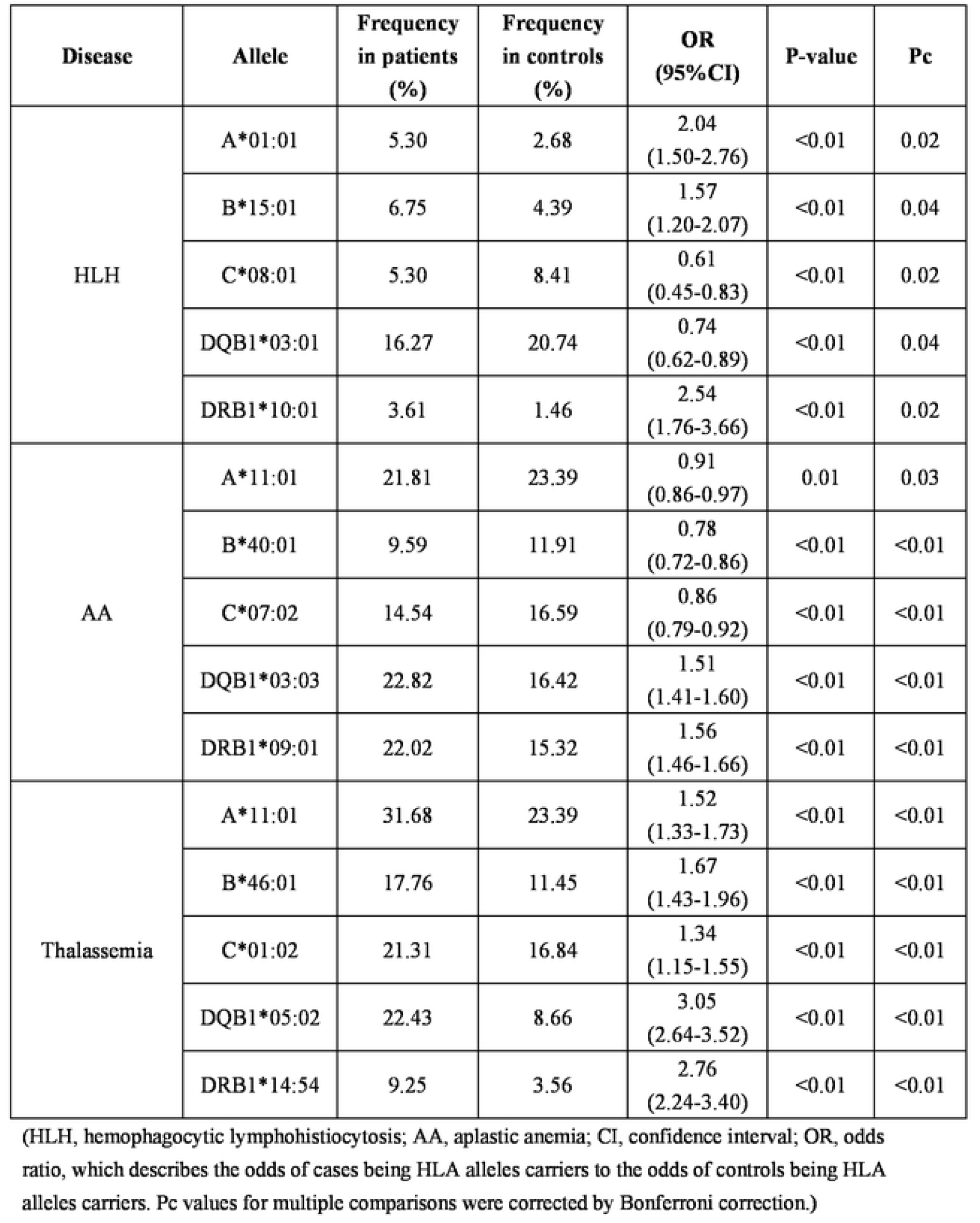
Differences in the most common HLA-A, -B, -C, -DQ, and -DR alleles between patients with benign hematologic disorders and controls.

The protective alleles for AA were mostly at HLA-DQB1 locus (10/32), with the highest frequency of A*11:01 (OR: 0.91; 95% CI: 0.86-0.97, Pc = 0.03), B*40:01 (OR: 0.78; 95% CI: 0.72-0.86, Pc < 0.01), C*07:02 (OR: 0.86; 95% CI: 0.79-0.92, Pc < 0.01), and DQB1*03:01 (OR: 0.90; 95% CI: 0.84-0.96, Pc < 0.01). HLA alleles at loci B and C accounted for more than half of the risk factors for AA (8/26 and 7/26, respectively); interestingly, the top two high-frequency risk alleles all belonged to class II [i.e. DQB1*03:03 (OR: 1.51; 95% CI: 1.41-1.60, Pc < 0.01), DRB1*09:01 (OR: 1.56; 95% CI: 1.46-1.66, Pc < 0.01).

### 3.3 Potential protective and risk HLA-A, -B, -C, -DQB1, and -DRB1 genotypes

We detected a total of 220 HLA-A-A genotypes, 198 HLA-B-B genotypes, 225 HLA-C-C genotypes, 182 HLA-DQB1-DQB1 genotypes, and 210 HLA-DRB1-DRB1 genotypes. Unfortunately, no risk/protective genotypes were found to be associated with malignant hematological diseases. But we observed the following susceptibility and antagonistic genotypes for benign hematological disorders.

Herein, 27 HLA genotypes seems to be protective factors for AA, including 8 of HLA-A, 7 of HLA-B, 3 of HLA-C, 3 of HLA-DQB1 and 6 of HLA-DRB1, with the more frequent ones of A*11:01-A*24:02 (OR: 0.70; 95% CI: 0.59-0.82, Pc < 0.01), C*01:02-C*07:02 (OR: 0.80; 95% CI: 0.67-0.95, Pc = 0.03), and DQB1*03:01-DQB1*06:01 (OR: 0.67; 95% CI: 0.54-0.83, Pc < 0.01). Whereas B*46:01-B*46:01 (OR: 1.67; 95% CI: 1.32-2.12, Pc < 0.01) and DRB1*09:01-DRB1*15:01 (OR: 1.77; 95% CI: 1.51-2.08, Pc < 0.01) occurred most frequently in the 25 risk genotypes for AA (**Table 3**).

**Table 3.** Variations in the most common HLA-A-A, HLA-B-B, HLA-C-C, HLA-DQ-DQ, and HLA-DR-DR genotypes in the benign hematopoietic patients and controls.

| Disease | Genotype | Frequency in patients (%) | Frequency in controls (%) | OR (95%CI) | P-value | Pc |
| --- | --- | --- | --- | --- | --- | --- |
| AA | A*11:01-A*24:02 | 5.20 | 7.30 | 0.70<br>(0.59-0.82) | <0.01 | <0.01 |
|  | B*46:01-B*46:01 | 2.58 | 1.56 | 1.67<br>(1.32-2.12) | <0.01 | <0.01 |
|  | C*01:02-C*07:02 | 4.71 | 5.83 | 0.80<br>(0.67-0.95) | 0.01 | 0.03 |
|  | DQB1*03:03-DQ<br>B1*06:01 | 8.12 | 3.49 | 2.44<br>(2.12-2.81) | <0.01 | <0.01 |
|  | DRB1*09:01-DR<br>B1*15:01 | 5.79 | 3.35 | 1.77<br>(1.51-2.08) | <0.01 | <0.01 |
| Thalassemia | A*11:01-A*11:01 | 10.84 | 6.03 | 1.89<br>(1.44-2.49) | <0.01 | <0.01 |
|  | B*46:01-B*58:01 | 3.74 | 1.64 | 2.32<br>(1.48-3.65) | <0.01 | <0.01 |
|  | C*01:02-C*03:04 | 7.66 | 3.83 | 2.08<br>(1.51-2.87) | <0.01 | <0.01 |
|  | DQB1*03:01-DQ<br>B1*05:02 | 7.66 | 3.41 | 2.35<br>(1.70-3.24) | <0.01 | <0.01 |
|  | DRB1*14:54-DR<br>B1*15:01 | 3.18 | 0.84 | 3.87<br>(2.38-6.32) | <0.01 | <0.01 |

For thalassemia, a statistically significant difference in HLA genotypes between patients and controls was observed, with only one protective genotype A*02:01-A*24:02 (OR: 0.36; 95% CI: 0.17-0.76, Pc = 0.03). A total of 22 risk genotypes of thalassemia consisted of 13 class I (5 for A, 6 for B, and 2 for C) and 9 class II genotypes (5 and 4 for DQB1 and DRB1, respectively), with B*46:01-B*58:01 (OR: 2.32; 95% CI: 1.48-3.65, Pc = 0.01), C*01:02-C*03:04 (OR: 2.08; 95% CI: 1.51-2.87, Pc < 0.01), DQB1*03:01-DQB1*05:02 (OR: 2.35; 95% CI: 1.70-3.24, Pc < 0.01), and DRB1*14:54-DRB1*15:01 (OR: 3.87; 95% CI: 2.38-6.32, Pc < 0.01) occurred most frequently (**Table 3**).

## 4 DISCUSSION

As the first large-scale HLA individual study in China, it focuses on hematologic diseases. The highlight is the exploration of differences in multiple hematologic disease types and HLA genes. In this study, alleles and genotypes of HLA-A, -B, -C, -DQ, and -DR were collected from 43,568 patients and 80,345 individuals without known pathology. This approach has revealed over 100 disease-associated loci (28 HLA-A, 37 HLA-B, 28 HLA-C, 21 HLA-DQB1, and 24 HLA-DRB1) and has provided insights into the hematologic disease surveillance **(Table S1-S7)**.

However, some high-frequency alleles and genotypes were common in both patients and individuals without known pathology. Notably, in malignant hematologic diseases, 3 high-frequency alleles (DQB1*03:01, DQB1*06:02, DRB1*15:01) are risky. For benign hematologic diseases, we observed 7 high-frequency alleles (A*01:01, B*46:01, C*01:02, DQB1*03:03, DQB1*05:02, DRB1*09:01, DRB1*14:54), and 3 high-frequency genotypes (A*11:01-A*11:01, B*46:01-B*58:01, B*46:01-B*46:01, C*01:02-C*03:04, DQB1*03:01-DQB1*05:02, DQB1*03:03-DQB1*06:01, DRB1*09:01-DRB1*15:01, DRB1*14:54-DRB1*15:01) are considered as risk genes. Thus these alleles and genotypes should be paid more attention.

For malignant hematologic diseases, Qi et al.(Qi *et al*. 2017) reported a higher frequency of DRB1*15:01 in Chinese ALL patients compared to controls as a risk allele (OR: 1.70; 95% CI: 1.24-2.33), and higher frequency of A*02:07 (Pc = 0.013), A*29:01 (Pc = 0.044), B*07:02 (Pc = 0.029), B*07:05:01G (Pc = 0.044) and B*35:02 (Pc = 0.023), but a lower frequency of A*02:03 in AML patients compared to controls (0.79% vs 3.10%, Pc = 0.011). Also, DRB1*11:28 occurred at a significantly higher frequency in the CML group compared with the controls (Pc = 0.015). Actually, our study further expanded the HLA alleles of AML and ALL susceptibility by increasing the sample size based on previous studies. Furthermore, Xiao et al.(Xiao *et al*. 2010) reported that the frequency of the HLA-DR15 expression in MDS patients (38.7%) was significantly higher than that in healthy controls (Pc < 0.01). The existence of the HLA-DR15 allele suggested MDS susceptibility, which is consistent with our findings. Uçar et al.(Uçar *et al*. 2016) discovered higher frequencies of HLA-A*29 (OR: 5.65; Pc = 0.001), B*07 (OR: 3.00; Pc = 0.003), and DRB1*11 (OR: 1.80; Pc = 0.002) alleles in patients with Hodgkin’s lymphoma compared with controls. Significantly increased allele frequencies of HLA-DQA1*01:03 and HLA-DQB1*06:01 have also been reported in lymphoma(Kawahara *et al*. 2005). Undoubtedly, malignant hematological diseases are mainly because of the occurrence of molecular abnormalities leading to the deregulation of immature hematopoietic cells(Dufva *et al*. 2020). Therefore, HLA loci are expected to be a novel biomarker for the clinical diagnosis of malignant hematologic disorders.

By comparing the results from previous research, we hope to determine HLA-related genes for benign hematologic diseases. However, there are no studies on the association of HLH with HLA genes in recent years, it must be pointed out we are the most comprehensive study of the Chinese population. Relatively, the association between HLA class II and anemia (especially, AA) has been reported more frequently. Yari et al.(Yari *et al*. 2008) reported DRB1*07 at a frequency of 8.3% in normal subjects and 15.7% in AA patients. Notably, Qi et al.(Qi *et al*. 2020) reported HLA-DRB1*15:01 (OR: 2.11; Pc = 2.87×10_-3_) and HLA-DQB1*06:02 (OR: 2.01; Pc = 1.86×10_-2_) were risk alleles of AA in Han population from northern China, which consistent with our findings. They also found HLA-DRB1*15:01-DQB1*06:02 (OR: 2.09; Pc = 4.90×10_-3_) and HLA-DRB1*14:05-DQB1*05:03 (OR: 2.82; Pc = 2.65×10_-2_) were strongly associated with AA. Therefore, it can be speculated that the diversity of HLA may be responsible for the differences between these studies. In addition, in a study of thalassemia patients in Iranian, HLA-DRB1*15:03 allele frequency was significantly different between 59 alloimmunised and 205 non-alloimmunised patients (OR: 4.193; Pc = 0.0001)(Darvishi *et al*. 2019). These preliminary data lay the foundation for further research of HLA in benign hematological diseases. Collectively, susceptible HLA alleles and genotypes may be considered a promising aspect of clinical surveillance.

There are several limitations to our study. First of all, the main limitation is the lack of diagnostic information for some patients, but our sample size is relatively sufficient compared to previous studies. Considering the errors caused by multiple comparisons, we used Bonferroni’s correction to adjust. Nonetheless, it can lead to overly conservative results when the number of multiple tests exceeds 10(Armstrong 2014; Sedgwick 2014). In addition, there is no consensus on the role of risk alleles, such as chromosomal abnormalities and other clonal evolution. Actually, some of our findings appeared inconsistent with previous reports. This possibly is due to deviations in cohort size and ethnic and regional distribution.

## 5 CONCLUSIONS

In summary, our results indicate several hematology-related HLA alleles and genotypes in the Chinese population. This contributes to the elucidation of the biological functions of related hematologic diseases, thus opening a new avenue for targeted immunopathologic therapy development. Ideally, these findings should be replicated in a study of regional distribution. And more attention should be paid to prognostic indicators (age, infection, medical, and family history). Despite the limitations, these are valuable in light of bringing further insight into HLA polymorphism and clinical transplantation of the Chinese population.

## Data Availability

All relevant data are within the manuscript and its Supporting Information files

## ACKNOWLEDGEMENT

We thank all the patients and clinical collaborators for participation in the study.

## STATEMENT OF ETHICS

This study was approved by the Ethics Committee of Shanghai Tissuebank Medical Laboratory, and all patients provided signed informed consent.

## AUTHOR CONTRIBUTIONS

Zhi-Yang Yuan, Zheng-Zhong Zheng and Ke-Ming Du designed the study, Yu-Xia Li performed the statistical analysis, Lin An and Zhong-Liang Liu contributed to clinical data collection, Ye-Mo Li and Dai-Yang Li, wrote the manuscript, Zheng-Zhong Zheng and Ke-Ming Du provided scientific advice and supervision. All authors have approved the final version of the manuscript.

## DATA AVAILABILITY

The data that support the findings of this study are available from the corresponding author, upon reasonable request.

## ETHICS STATEMENT

This study protocol was reviewed and approved by the ethics committee of the Shanghai Tissuebank Medical Laboratory, approval number 2020-006.

